# Mobility and COVID-19 in Andorra: Country-scale analysis of high-resolution mobility patterns and infection spread

**DOI:** 10.1101/2021.02.18.21251977

**Authors:** Ronan Doorley, Alex Berke, Ariel Noyman, Luis Alonso, Josep Ribó, Vanesa Arroyo, Marc Pons, Kent Larson

**Author notes:** Authors RD, AB, AN, LA and KL are with the City Science Group of the MIT Media Lab. Authors JR, VA and MP are with the Andorra Innovation Hub.

## Abstract

In the absence of effective vaccines, non-pharmaceutical interventions, such as mobility restrictions, were globally adopted as critically important strategies for curbing the spread of COVID-19. However, such interventions come with immense social and economic costs and the relative effectiveness of different mobility restrictions are not well understood. This study analyzed uniquely comprehensive datasets for the entirety of a small country, consisting of serology data, telecoms data, and COVID-19 case reports, in order to examine the relationship between mobility and transmission of COVID-19.

Andorra is a small European country where tourism is a large part of the economy. Stringent mobility restrictions were put in place in Spring 2020. Additionally, 91% of the population participated in a voluntary COVID-19 serology testing programme and those data were made available for this study. Furthermore, high resolution telecoms data for the entire population were available for analysis of mobility and proximity patterns. A set of mobility metrics were developed to indicate levels of crowding, stay-at-home rates, trip-making and contact with tourists. Mobility metrics were compared to infection rates across communities and transmission rate over time.

Several of these metrics were highly correlated with transmission rate, with a lead time of approximately 18 days, with some metrics more highly correlated than others. There was a stronger correlation for measures of crowding and inter-community trip-making, and a weaker correlation for total trips (including intra-community trips) and stay-at-homes rates.

## 1 Introduction

The rapid spread of the COVID-19 pandemic in 2020, has led to an unprecedented global effort to curb the disease and its resulting mortality and morbidity. As of early 2021, given the lack of effective therapeutics or widely distributed vaccines, Nonpharmaceutical Interventions (NPIs) remain the primary public health strategies for achieving these goals [10, 23]. While several COVID-19 vaccines are being offered to European and global populations in early 2021, it might take significant time for vaccinations to accomplish ‘herd immunity’ and thereby significantly affect the trajectory of the pandemic. Furthermore, many countries in the developing world will likely not reach adequate levels of vaccination until 2022 or later [38]. It is therefore fair to asses that along-side vaccination efforts, NPIs will continue to be an important tool in the fight against COVID-19 for the foreseeable future.

During the pandemic, government mandates restricting mobility and economic activity have become widespread in ways that were unimagined only months previously. Economic lockdowns, border closures and mobility restrictions are clearly effective to some degree in curbing transmission of COVID-19 [6]. However, these measures have enormous economic, social and mental health impacts and therefore policy-makers must carefully consider the relative costs and benefits of every policy. Unfortunately, neither the costs nor benefits of NPIs for COVID-19 are well understood due the enormous complexity of the problem in terms of both virology and human dynamics. To further complicate matters, governments have been forced to introduce wide ranges of measures at once, from closing borders to restricting people’s radius of movement. This has made it difficult to directly study the impacts of any one intervention on transmission rates.

Until recently, gathering fine grained movement patterns data for large populations of people was nearly impossible [31]. With the advent of GPS-enabled smartphones, most people around the world today are continuously generating data on their mobility patterns. These data are collected and aggregated in several ways: Telecoms companies routinely record histories of device connections to cell towers for billing purposes, and these data can be used to reconstruct individual device trajectories. There are also companies who collect location data from users through Software Development Kits (SDKs) embedded in a set of partner apps [11, 32]. These data can have finer spatial resolution than those collected by telecoms companies due to the use of GPS. However, since they only collect data from users who use specific apps and opt in to having their location data collected, sample sizes are small and there is potential for demographic self-selection bias.

Numerous studies that use mobile phone data to analyze urban mobility and behaviour have been published over the past several years [20, 21, 31]. In the past year, there have also been several studies using telecoms data to study mobility in the context of the COVID-19 pandemic. Some studies only look at how behaviour has changed and do not incorporate data on COVID-19 infections [7, 9, 24, 32]. Other studies build metapopulation compartmental models, which divide the population into smaller communities and assume that transmission within each community is governed by a model such as random mixing, with reduced transmission between communities [25]. Previously, the mixing of communities in these models has been informed by traditional commuting data but recent studies have used O-D matrices and/or visits to POIs inferred from telecoms data [5, 39]. As discussed in [22], this approach assumes a known model of the exact social-contact pattern of individuals within each origin, destination and/or POI. The sub-communities should therefore be small enough that contact patterns are uniform, and the model for these contact patterns must be realistic. The most commonly assumed model of contact patterns is mass-action, whereby interactions per person are independent of the density of people. However, other studies assume that infection rates in a location are proportional to density of people [12]. There is also variation across recent studies in the size of the area used as sub-communities. The accuracy of such simulation models depend on a good understanding of the influence of density on transmission within communities.

This study is based in the country of Andorra and explores the relationships between government policies, several inferred mobility metrics and the nature by which COVID-19 spread across the country. More specifically, this work attempts to answer the following questions: How did travel patterns change before, during and after lockdown policies were implemented? Did changes in mobility behaviour and crowding over time correspond to changes in transmission rate over time? Which changes in mobility behavior are most closely correlated with changes in transmission rate?

The study makes use of telecoms data for all mobile users in the country for a period before and during the government lockdown, allowing for a comprehensive analysis of the population’s mobility behaviors over time. Additionally, this study makes use of COVID-19 antibody test results covering over 91% of the population. The combination of full coverage telecoms data and antibodies data provided a unique opportunity to study the spread of COVID-19 in a relatively closed population.

Section 2 provides context for the work by briefly introducing the COVID-19 pandemic in Andorra. The data sources, pre-processing and mobility metrics used in the study are then described in section 3. Section 4 presents analysis and findings about the relationship between mobility and COVID-19 infections in Andorra. The implications of the findings are discussed in section 5, followed by a conclusion.

## 2 Andorra and COVID-19

The study region for this work was the European microstate of Andorra. Andorra is located in the heart of the Pyrenees mountains, situated between the borders of France and Spain. It lacks an airport or train service so the primary way to enter or exit the country is by crossing the French or Spanish border by car (see Figure 1). For this reason, the stringent border closures by France and Spain during the pandemic effectively closed the borders of Andorra.

**Figure 1:**
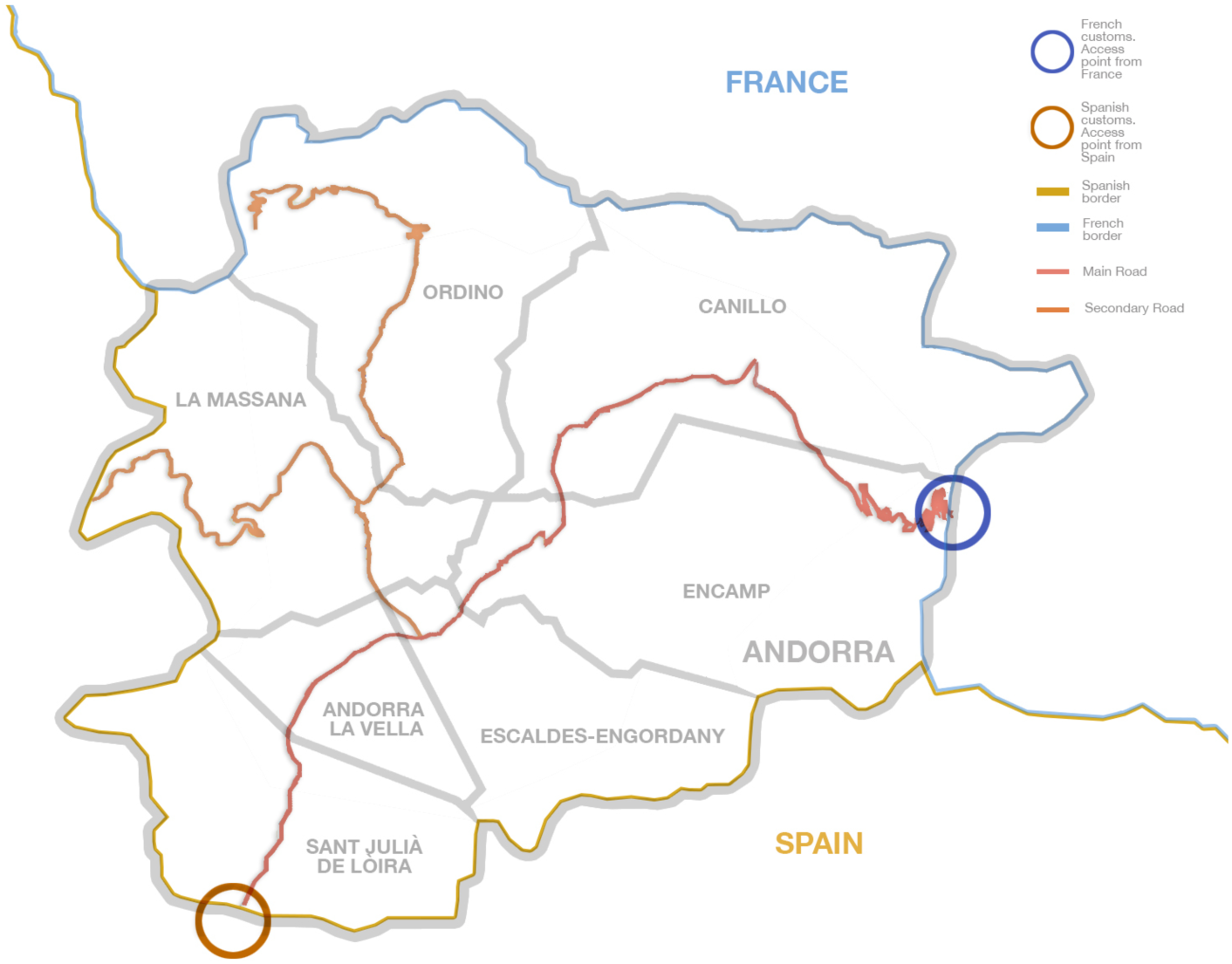
Andorra is situated between France (north) and Spain (south). It has no airport or train service so the primary means to enter or exit the country is by crossing the French or Spanish borders by car. Highlighted are the main entry routes to the country, as well secondary roads leading to the different parishes.

Andorra is small in terms of both geography and population, and derives much of its income from the tourism industry. It has a population of approximately 77,000 people and receives about 8 million visitors during normal seasons [13]. Partly due to Andorra’s small size, the government is able to collect and provide comprehensive datasets that contributed to our study. There is one telecoms provider for the entire country, which contributes a comprehensive view of all mobile subscribers who spend any time in Andorra, whether they are Andorran nationals or have foreign SIM cards. Andorra was also able to conduct serology tests for its entire population (91% participation rate). Moreover, Andorra was able to implement swift and comprehensive policy measures at the start of the pandemic. The ability for Andorra to implement new and comprehensive policy changes will likely continue with future developments in the epidemic. On March 2, 2020, the country’s first coronavirus case was confirmed. Announcements of lockdown measures beginning on March 13, as well as border closures [14,30], soon followed. A list of key dates and policies is given in Table 1.

**Table 1:** Timeline of the COVID-19 related dates in Andorra

|  |  |
| --- | --- |
| March 2 | First coronavirus case confirmed in Andorra. |
| March 13 | Partial lockdown by Andorran government. |
| March 17 | French border closure. |
| March 18 | Total lockdown by Andorran government. |
| March 19 | Spanish border closure. |
| April 7 | Beginning of masks delivery and progressive use by population. |
| April 17 | Allowance of 1 hour of walk in 1km radius every two days. |
| April 20 | Phase 1 reopening. Low risk economic activities resume. 1000 people return to work. |
| May 1 | Start of 1st round of population serology screening. |
| May 4 | Phase 2 reopening. Additional 4760 workers return to normal activity. |
| May 13 | Increase to 2 hours of activity every day to walk or exercise. |
| May 14 | Start of 2nd round of population serology screening. |
| May 18 | Phase 3 reopening. Additional 3300 workers return to normal activity. |
| June 1 | Confinement restrictions completely lifted. |
| June 15 | French and Spanish borders are open (with some restrictions on the Spanish side). |
| June 21 | Neighboring country, Spain, lifts state of emergency and lifts some border controls. |
| July 1 | Remaining Andorran restrictions on Spanish border are lifted. |
| Sept. 1 | Massive testing for teachers and school children. |
| Oct. 7 | Bars and restaurants access is restricted. |
| Oct. 10 - 12 | Massive entry of Spanish tourists due to Spanish holiday. |
| Oct. 22 | Massive testing strategy to test 40% of the population every week begins. |
| Oct. 26 | Restrictions on movement between both the Spanish and French borders. |
| Oct. 28 | Restrictions on in-person meetings. Working at home is mandatory if possible. |

## 3 Data and Methodology

Three primary data sources were used in this study: (i) the results of COVID-19 serology testing in which more than 91% of the population participated, (ii) daily reported COVID-19 case counts for Andorra and (iii) the telecoms data for nearly all subscribers within Andorra. The telecoms data were used to develop and analyze metrics that are indicative of people’s mobility and behaviors. The mobility and behavioral metrics include the daily number of entrances to the country, the fraction of the population staying home, the number of people making trips between the country’s regions (parishes), and measures of crowding. In addition, serology and reported cases data were used to analyze infection rates and the temporal relationship between infection rates and mobility behaviors. Each of the data sources and the computation of metrics are described in the sub-sections below.

### 3.1 Reported Cases Data

This analysis uses daily confirmed COVID-19 cases reported from March 1st to October 31st of 2020. This period spans the time from the first reported case in Andorra to the end of the period of mobility metrics used in this study. The data source is the COVID-19 Data Repository by the Center for Systems Science and Engineering (CSSE) at Johns Hopkins University [19].

The reported cases data are subject to error due to reporting lags and irregular testing periods. In particular, there was an influx of late case reports on June 3rd which were identified through the mass serology testing. These reports were removed for the purposes of temporal analysis because they were not registered at the actual date of testing, and because they resulted from testing which was not carried out at other time periods and would therefore bias the temporal analysis. To mitigate reporting irregularities, this analysis smooths the case reports data by averaging over a rolling window of 14 days (see Figure 2).

**Figure 2:**
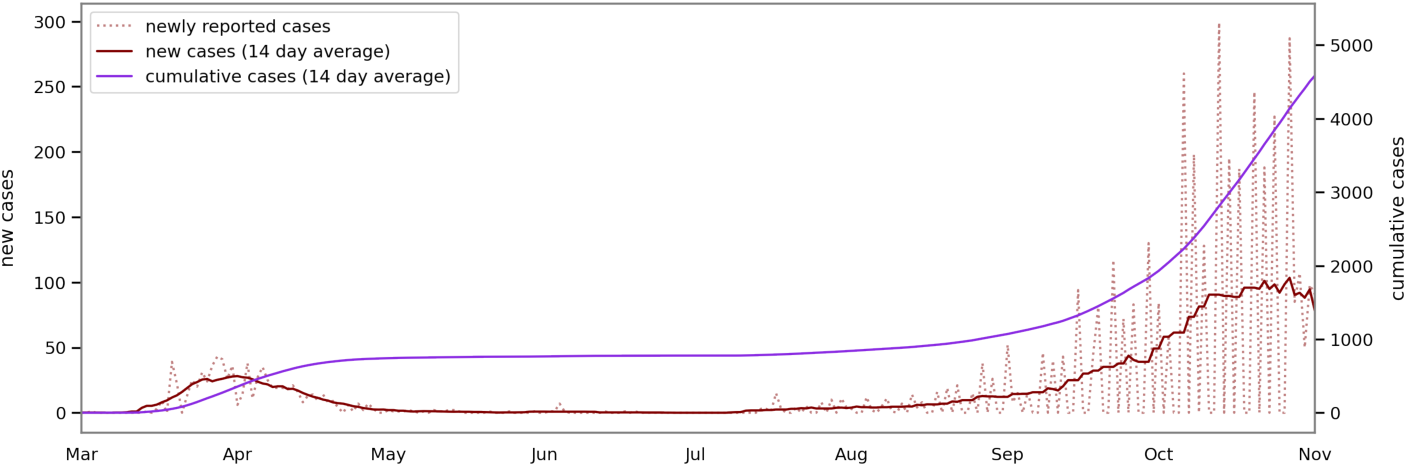
A timeline of new and cumulative case reports. The dotted line shows the day cases are reported. The solid lines show values smoothed over a 14-day rolling average.

### 3.2 Serology Data

In May, 2020, the Andorran government launched a campaign to provide voluntary serology testing for its entire population. The screening was conducted for a previous research study [36], which was approved by the relevant Andorran agencies and local research ethics committee. As shown in Figure 2, during this period and the preceding weeks, the cumulative case count was relatively flat, implying that the number of people with either current or prior infections remained stable during this period. Participants were tested for presence of both IgM and IgG antibodies twice, with two weeks between each round of testing. 91% of the population completed the first round of testing and 86% completed the second round. The test data were annonymised, so that individual-level analysis of risk was not possible. However, the data included additional variables, including parish of residence and residency status, allowing group infection rates to be compared. Livzon IgM/IgG Diagnostic Kits were used [29]. All such screenings are subject to some level of error, both false positives and false negatives. Independent testing of the kits had previously been carried out to establish the sensitivity and specificity of the tests. Maximum likelihood estimation was used to estimate the actual number of people with current or previous infections, accounting for sensitivity and specificity [35], as shown in equation 1 below. Having estimated the overall prevalence, the probability of every individual having a current or previous infection was calculated using Bayes’ Rule as shown in equation 2.

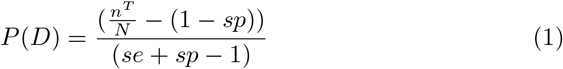

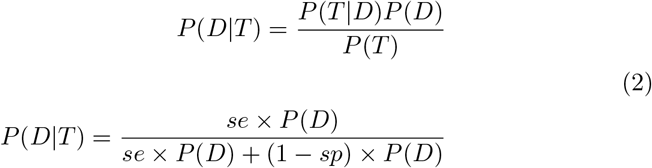

Where

*n*^*T*^ = number of positive tests

*N* = size of the population

*D* = actually has a current or previous infection

*T* = tested positive for IgG and/or IgM antibodies

*se* = combined sensitivity of the test for IgG and/or IgM

*sp* = combined specificity of the test for IgG and/or IgM

### 3.3 Telecoms Data

For this study, telecoms data were provided by Andorra Telecom (AT). Since AT is the sole telecoms provider in Andorra, the data cover all mobile subscribers in the country, unlike most telecoms datsets where the market is fragmented. Each observation in the AT data includes a unique ID for the subscriber, a timestamp, the coordinates of the device at the time of the update, and nationality for the subscriber’s home network. The coordinates are given in the standard WGS84 system and estimated by AT to be accurate to within 25-100m in urban areas. AT utilizes enhanced triangulation using the angle, time and RSS received by the device from different base-stations as well as error mitigation using a proprietary algorithm. The AT data have been further described in [20, 31]. In order to comply with data protection policies, the telecoms data were anonymised and only used to compute metrics which contained no identifying information. Any attributes associated with metrics, such as parish of residence, represent groups of several thousand people at least.

This study used data from March 2019 to June 2019 and January 2020 to October 2020, from both the 3G and 4G networks. Data recorded by AT were missing or incomplete in periods of the study (February 14 - March 1, June 28 - June 29, July 21 - July 27, and October 1 - October 6, 2020). These gaps resulted in missing values in the computed mobility metrics, and are displayed as gaps in the time series plots of these metrics.

#### 3.3.1 Pre-Processing

In 2020, the population of Andorra was approximately 77,000 people [18]. During the period of January through October 2020, a total of 1,288,799 unique subscriber IDs were observed in the AT data. The vast majority of these subscribers were tourists who entered the country with both Andorrans and international SIM cards. As described below, the data for all subscribers were pre-processed in order to identify stay-points, home parishes, days present in Andorra and residency status.

To reduce noise, and increase the probability of actual location, the series of location observations for each subscriber was reduced to a series of stay-points of 10 minutes or more within a radius of 200m or less. The stay-point extraction algorithm of Li et al. (2008) [28] was used for this purpose. In order to compute daily mobility metrics across all people present in the country, it was necessary to know which people were present on any day. Some devices were found to be unobserved in the data for several days, even during the full government lockdown. This may be due to a combination of inactivity, lack of telecoms reception in certain areas, and/or noisy data. It was assumed that gaps in data of two weeks or more represented true absence from the country. The beginnings and endings of periods of presence were counted as entrances to and departures from the country respectively.

By observing the distribution of total number of days present across all subscribers, it was found that the majority of subscribers were present fewer than 50 days. The rest of the subscribers were present for a significantly greater number of days. 50 days of presence in Andorra was therefore considered as an appropriate cutoff for identifying tourists (see Figure 7.14 in the Supplementary Material). Further analysis uses this categorization of tourists versus non-tourists.

##### Home parishes

Home parishes were inferred for each subscriber on a monthly basis, since some residents of Andorra moved during the periods of study and since the tourist population fluctuated. To infer home parish, each stay-point first was assigned to the parish in which it was contained. Each subscriber’s home parish was then determined to be the parish in which they spent the most cumulative time during night-time hours (12:00am to 6:00am). This method is similar to those employed in related studies of human mobility that use cellular data [15, 26, 32, 33]. To evaluate the representativeness of the telecoms data and this methodology, the distribution of home parishes inferred for subscribers was compared to the projected 2020 population statistics published by the Andorran government [18]. This evaluation used AT data from May, 2020 and was restricted to the subscribers using Andorra as their home network, and that the pre-processing stage categorized as non-tourists. The month of May was used because it was the last month of the lockdown before the country reopened its borders. These data restrictions were made in order to more likely capture the permanent residents of Andorra for a better comparison to the published population statistics. The Pearson correlation coefficient between the parish-level populations inferred from this analysis and the published population statistics is 0.959, suggesting that the telecoms data is highly representative of the true population (see Figure 3).

**Figure 3:**
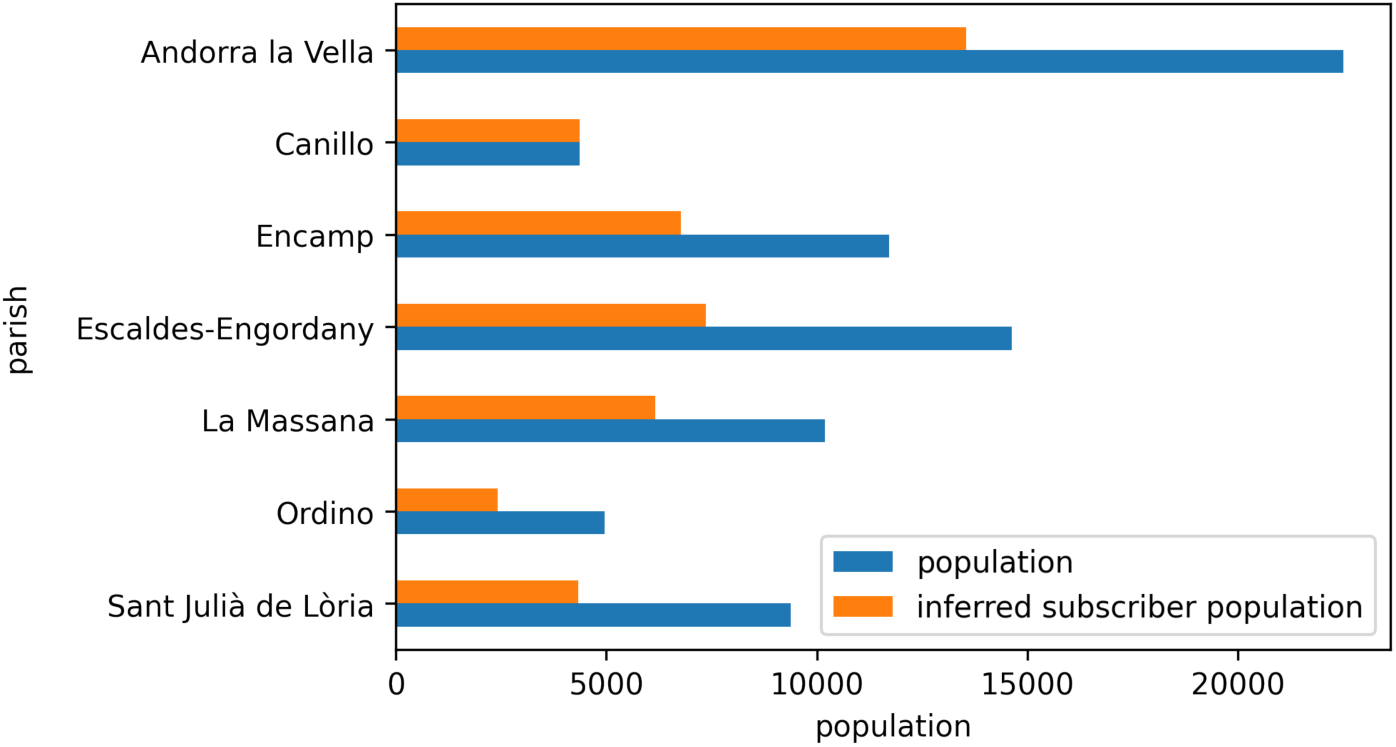
The parish-level populations inferred from the telecoms data for May 2020 were compared to the published 2020 population statistics [18]. There is a Pearson correlation coefficient of 0.959, suggesting that the telecoms data is highly representative of the true population.

#### 3.3.2 Mobility Metrics

A number of key mobility metrics were computed based on the AT data and used for this report. The metrics are described below.

##### Stay-at-Home Rates

Advising people to stay at home as much as possible is widely recognised as an effective policy for reducing transmission of infection. The stay-at-home rate for a parish on a given day was defined as the number of residents of that parish present and staying home divided by the total number of people present. A person was assumed to have stayed at home if they had at most one stay-point in their own parish and no stay points in other parishes. This assumes that anyone making a trip would have recorded at least two stay-points as they moved between cell coverage areas.

##### Trips

One purpose of lockdown policies was to reduce the population’s mobility, and the risk of a mobile population potentially carrying disease between places they go. The population’s mobility may be coarsely viewed by its trips between places. This analysis uses the number of total daily trips as well as the daily number of trips between parishes. Trips between parishes may be more indicative of when people make larger trips outside their community and so may also be associated with higher risk. This analysis further uses the daily number of distinct subscribers present in Andorra who are making any trips, as well as those making trips between parishes.

Daily trips for subscribers are counted as their daily number of stay points minus 1, since a new stay point is recorded when a subscriber moves beyond a 200m radius. Trips between parishes are counted when consecutive stay points occur in different parishes.

##### Interaction Potentia

Dense crowds of people create high potential for human interaction and therefore transmission. Indoor crowding involves the additional risk of aerosol transmission even when efforts are made to social distance. As a proxy for such crowding, interaction potential was defined as follows. The time period and study region were first divided into discrete spatio-temporal intervals. H3 cells of resolution 11 [1] were used as the geographic intervals. At this resolution, the H3 cells have an approximate diameter of 50m [4] which roughly corresponds to the resolution of the telecoms data in urban areas. Time was divided in 20 minute intervals. In every spatio-temporal interval, the number of potential interactions was counted as the number of unique person pairs in the interval. This is illustrated in Figure 4. The interactions were further subdivided by the home parishes and/or residence status of the people. Additionally, a distinction was made between indoor interaction potential and outdoor interaction potential based on the percentage of indoor and outdoor area in the H3 cell. These percentages were calculated using the Global Human Settlement Layers (GHSL) [16]. The interaction potential metrics can be defined as below:

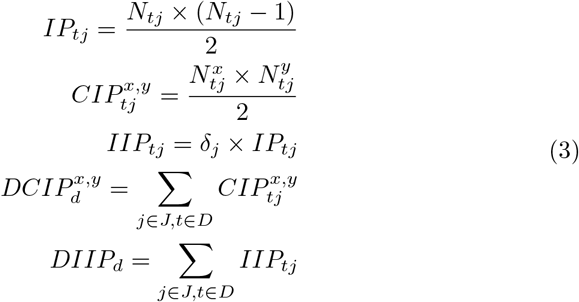

**Figure 4:**
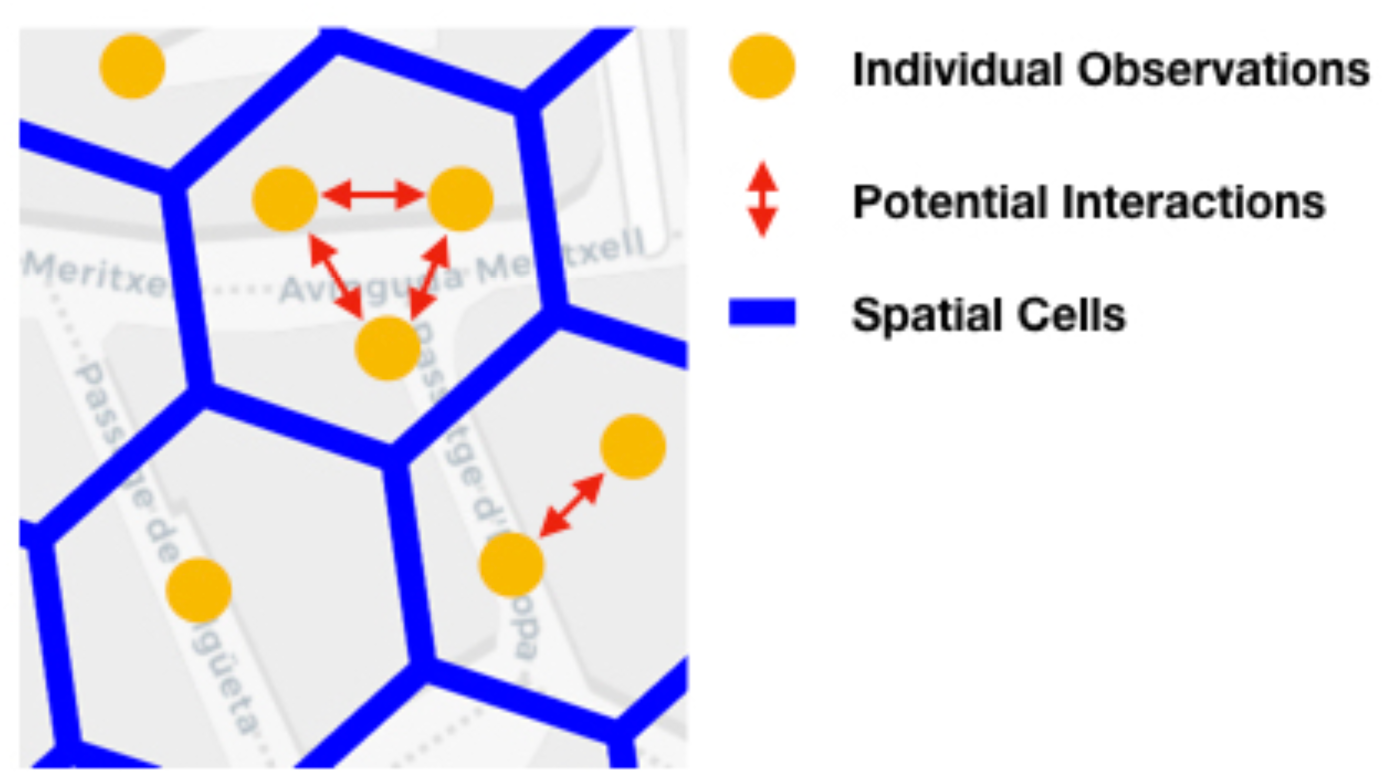
The computation of interaction potential based on individual spatial observations contained in H3 cells.

Where

*IP*_*tj*_ = Interaction Potential: the number of potential interactions in cell *j* during time interval *t*.

*N*_*tj*_ = the number of subscribers observed in cell *j* during time interval *t*.

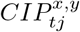= Cross Interaction Potential: the number of potential interactions between person categories *x* and *y* in cell *j* during time interval *t*.

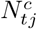= the number of subscribers of category *c* observed in cell *j* during time interval *t*.

*IIP*_*tj*_ = Indoor Interaction Potential: the number of potential interactions in cell *j* during time interval *t*, attributable to indoor areas.

*δ*_*j*_ = the fraction of land area of cell *j* covered by buildings.

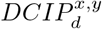= Daily Cross Interaction Potential: the number of potential interactions between person categories *x* and *y* during day *d*.

*DIIP*_*d*_ = Daily Indoor Interaction Potential: the number of potential interactions during day *d*, attributable to indoor areas.

*D* = the set of all intervals *t* in day *d*.

*P* = the set of all intervals *t* in the analysis period.

*J* = the set of all spatial cells in the study area.

Unique person-to-person interactions per day were also quantified. The daily person-to-person interaction graph, *G*_*d*_ was defined as follows. A node was created for every person present in the country that day. A pair of nodes was linked if those two people had been co-located in one or more spatio-temporal intervals during that day. The degree of each node was defined as the number of Daily Contacts of the corresponding person.

## 4 Analysis and Findings

### 4.1 Infections

In the first round of serology testing, IgM and IgG antibodies were detected in 9.6% of participants. In the second round of testing, this fell to 8%. In addition, participation rates fell from 91% to 86% between rounds. The reduction in antibody detection suggests that the antibodies fell below the detection limit rapidly. For this reason, as well as the reduction in test participation, the first round results alone were considered as a better indicator of overall exposure, while still being conservative. Using the maximum likelihood estimation formula of equation 1, the overall infection rate was estimated to be 11%. By comparing the number of reported cases before May 1st to the inferred number of infections, it was estimated that just 1 in 11 cases of COVID-19 were recorded in official statistics before this date.

The probabilities of each individual having a current or prior infection during the first round were then calculated using Bayes’ Rule as described in equation 2. These probabilities were then averaged across residence parishes and residency status to estimate the percentage of infections in each group. Note that the adjacent parishes of Andorra La Vella and Escaldes-Engordany were combined for the serology analysis. This was done on the advice of Andorran officials because the two parishes comprise a single urban area and were sometimes interchanged in the address field of the serology data. The populations and infection rates in each parish are shown in Table 2. The overall infection rate for temporary workers was estimated to be 14% compared to 11% for ordinary residents. La Massanna was the parish with the highest overall infection rate of 14% compared to the lowest rate of 9% in Andorra la Vella. In order to determine whether the differences in infection rate across temporary status and parish of residence were significant, chi squared tests were performed. It was found that both temporary status and parish of residence were significantly associated with infection rate (p*<*1e-5).

**Table 2:**
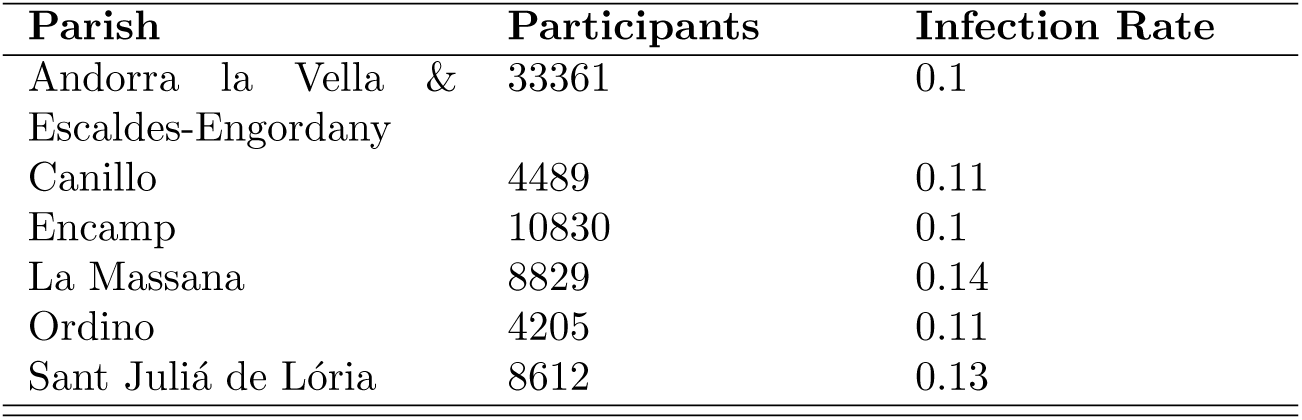
Participants numbers and aggregated infection rates by parish from serology testing.

| Parish | Participants | Infection Rate |
| --- | --- | --- |
| Andorra la Vella & Escaldes-Engordany | 33361 | 0.1 |
| Canillo | 4489 | 0.11 |
| Encamp | 10830 | 0.1 |
| La Massana | 8829 | 0.14 |
| Ordino | 4205 | 0.11 |
| Sant Julià de Lòria | 8612 | 0.13 |

### 4.2 Mobility

The mobility of the population is reported on in several phases. Firstly, in order to understand the initial outbreak, mobility patterns during January and February 2020 were analysed with an emphasis on tourists. Mobility patterns from March onward were then analyzed in order to assess how the population responded to the pandemic and to government policies. The distribution of mobility behaviours is also analyzed in order to explore whether highly active people and events may have disproportionately contributed to transmission risk. The evolution of mobility behaviour over time is then compared to the transmission rate during this period.

#### 4.2.1 Tourism before Outbreak

The pre-outbreak analysis was focused on the period from January 1st to February 14th, 2020. Unfortunately, the telecoms data for the last 2 weeks of February were unavailable. The first recorded case of COVID-19 in Andorra was on March 2. January and February are part of the ski season in Andorra and ski resorts were identified as high risk environments for the spread of COVID-19 in Europe [2, 3, 17]. Mobility of tourists and contact with tourists were therefore key considerations. The largest number of tourists during this period (45%) were found to have stayed in the parish of Canillo. In order to more directly assess the level of interactions between tourists and residents of each parish, people were categorised as either a resident of one of the seven parishes or as a tourist. Tourists were further categorised as Spanish, French or Other based on their their home network. The average Daily Cross Interaction Potential between each of these 10 groups was computed as described in section 3.3.2. As shown in Figure 5, the potential for interaction with tourists was much higher for residents of Canillo than for any other parish.

**Figure 5:**
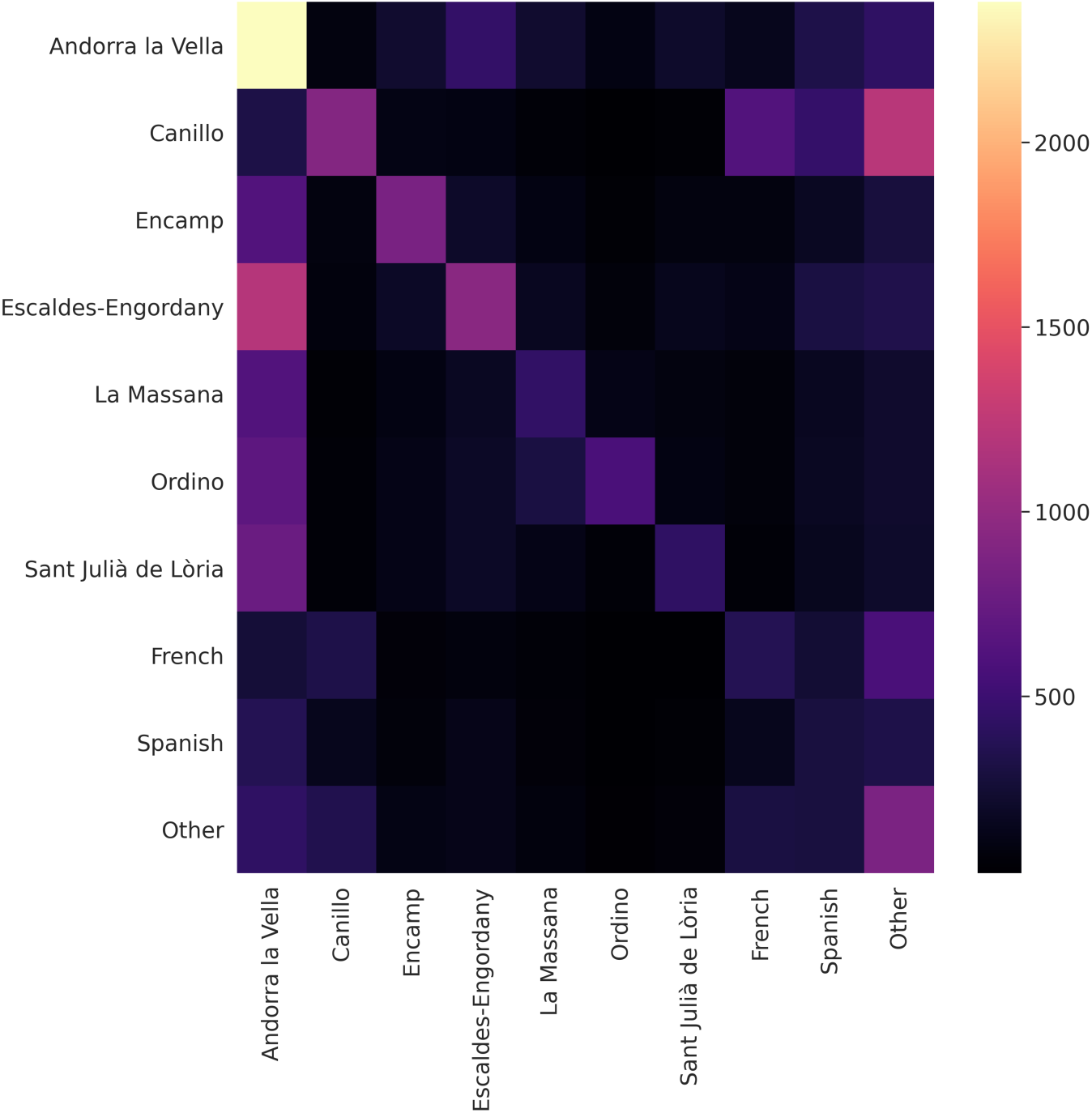
Average Daily Cross Interaction Potential (DCIP) between residents of each parish and tourists by country of origin. The value of row i, column j corresponds to the DCIP between groups i and j, normalised by the population of group i.

#### 4.2.2 Entrances and Departures

Throughout the pre-lockdown and lockdown periods, entrances into and departures from the country corresponded with border restrictions. As shown in Figure 6, this behavior differed by residency status. As expected, the initial lockdown policies that began March 13th, and the border closures by neighboring countries, led to mass departures from Andorra. This is observed in the telecoms data as a large drop-off in the number of subscribers present in mid-March. Many of these subscribers were tourists, and some may also have been temporary workers. The population was relatively stable between mid March and the end of May while lockdown was in effect and serology testing was carried out. In early May, there was a small surge of non-tourists arriving to the country and towards the end of May, there was a small surge of non-tourists leaving the country. This is shown in Figure 6. These observations may reflect people arriving in order to participate in serology testing as well as leaving the country after receiving one or both test results. Further analysis found that the surge in departures by non-tourists at the end of May was driven by people who did not return to the country.

**Figure 6:**
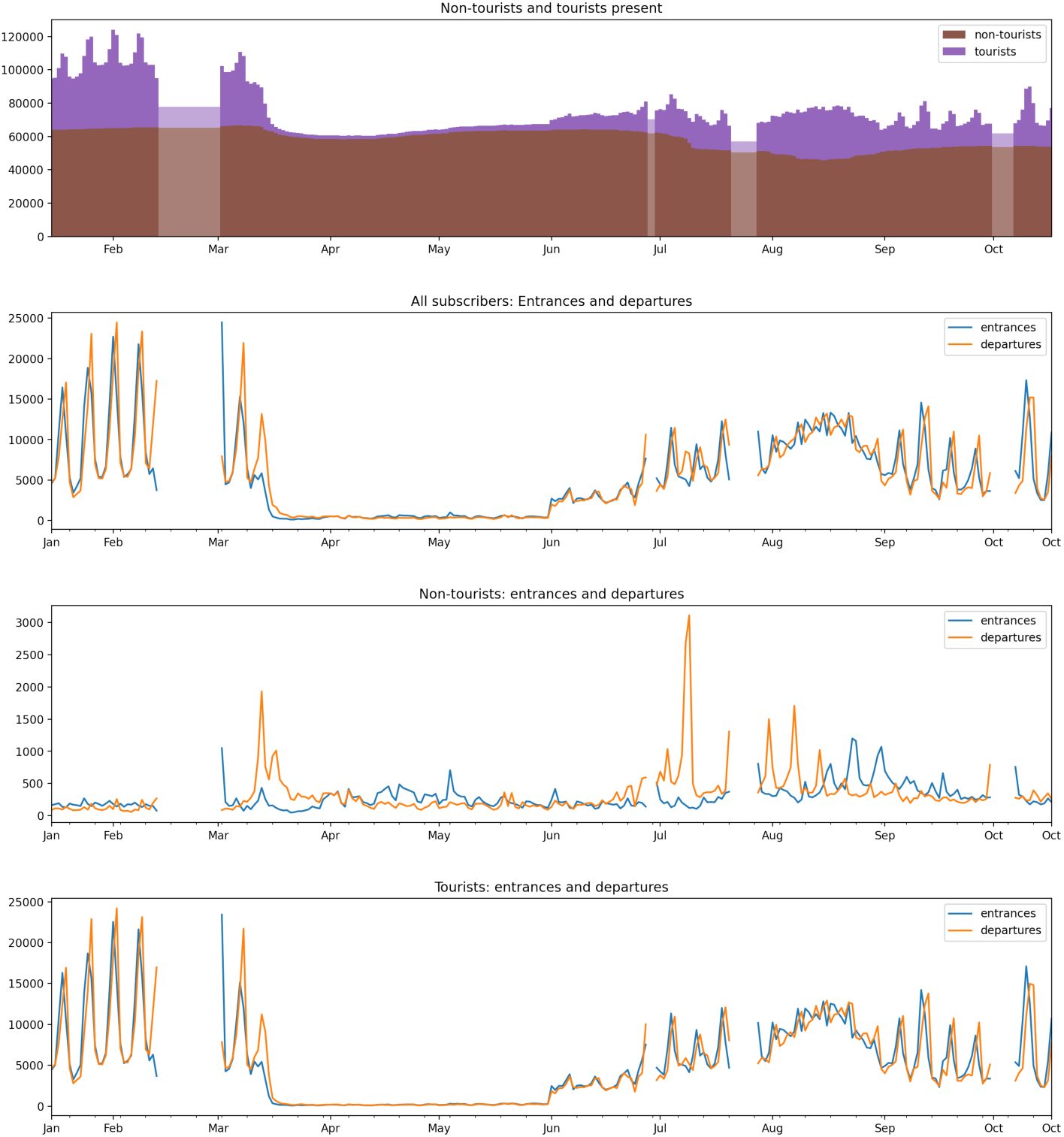
Daily presence, entrances, and departures inferred from telecoms data, by residency status (tourists versus non-tourists). There were gaps in the available telecoms data during the study (February 14 - March 1, June 28 - June 29, July 21 - July 27, and October 1 - October 6, 2020). These data gaps resulted in missing values in the computed mobility metrics, and are displayed as gaps in the time series plots of these metrics. The gaps also resulted in spikes in inferred departures and entrances directly before and after the gaps, respectively.

On June 1st, the lockdown in Andorra was lifted and both tourists and non-tourists immediately began to enter the country again. Initially, the tourists were mostly French, followed by an influx of Spanish tourists later in June, corresponding to the reopening of Spanish borders on June 21st. There was a spike in departures by non-tourists in the second weekend of July (July 9 saw over 3100 departures), corresponding to the weekend at the start of the common holiday period of Andorrans.

#### 4.2.3 Lockdown Mobility Behaviour by Parish

Daily mobility metrics were computed by parish in order to explore whether there were any differences in mobility behaviours or adherence to social distancing policies by parish and whether such differences corresponded to differences in infection rates by parish.

Andorra’s parishes differ in the extent to which the number of people within them are tourists in a normal year. The differences in their population characteristics elicit differences in their mobility behaviors. Differences in their tourism populations are displayed in Figure 7. It shows the daily number of Andorranm and non-Andorran subscribers reporting stays in each parish, comparing 2019, a normal tourism year, to the same period in 2020. In a normal tourism season, the populations of some smaller parishes, such as Canillo, are primarily non-Andorran. A mass departure by non-Andorran subscribers can be seen at the start of the lockdown in mid March 2020. In a normal year there would have been spikes in the non-Andorran tourist population throughout the spring, particularly on weekends. This influx of tourism did not occur in 2020, as it was prohibited by border restrictions that did not subside until June 1st.

The parishes differed in the extent to which their residents stayed at home during the lockdown, as shown in Figure 8. All parishes exhibit a similar trend with a sharp increase in the portion of people staying home at the start of the most restrictive lockdown measures in mid March. There is then a gradual drop off as lockdown policies were relaxed. At the start of the lockdown period, there were two distinct clusters of parishes in terms of the average stay-at-home rate: Andorra la Vella, Canillo, and Sant Juli’a de L’oria exhibited the highest portion of users staying home. However, this difference gradually diminished as the months continued.

**Figure 7:**
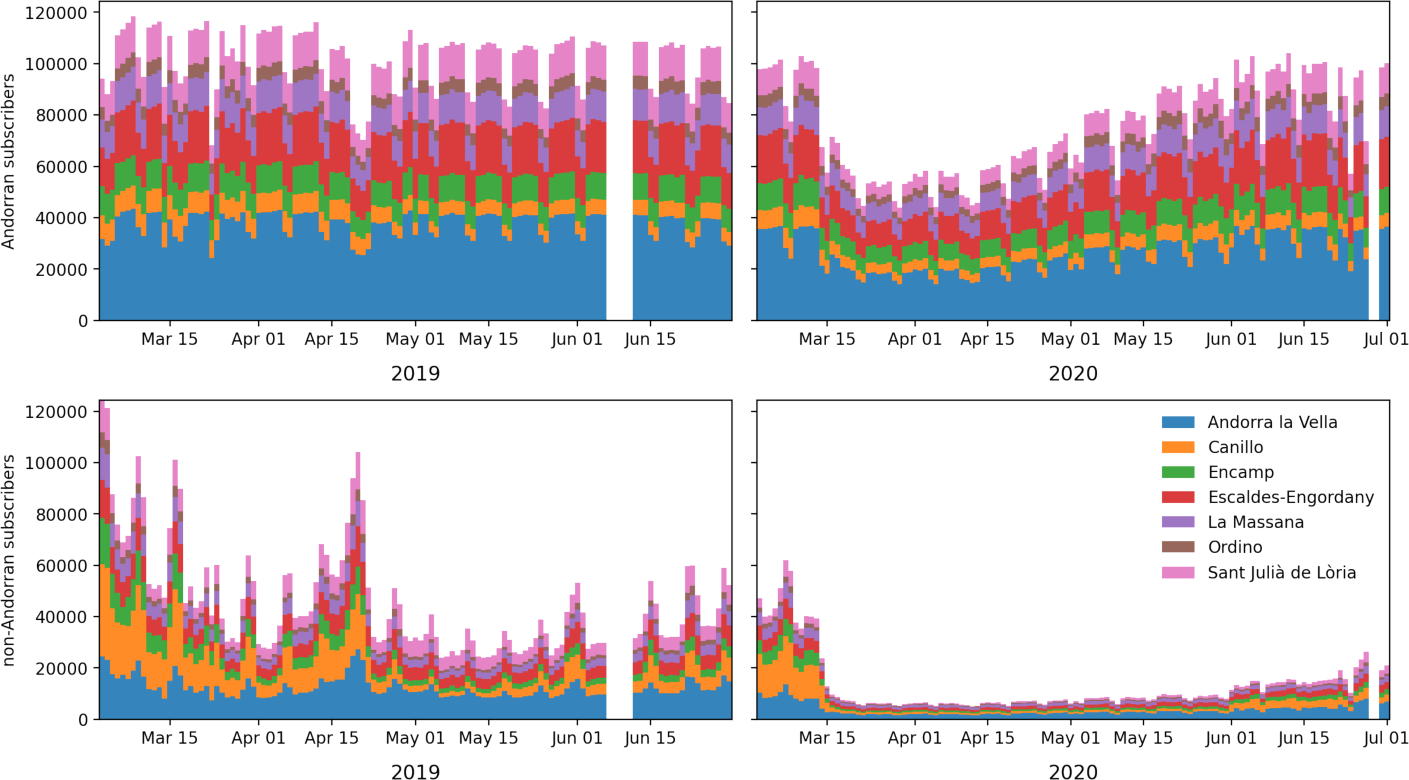
Daily subscribers reporting stays in each parish for March through June, comparing 2020 to 2019. A mass departure from the smaller tourism heavy parishes can be seen at the start of the lockdown in mid March 2020.

**Figure 8:**
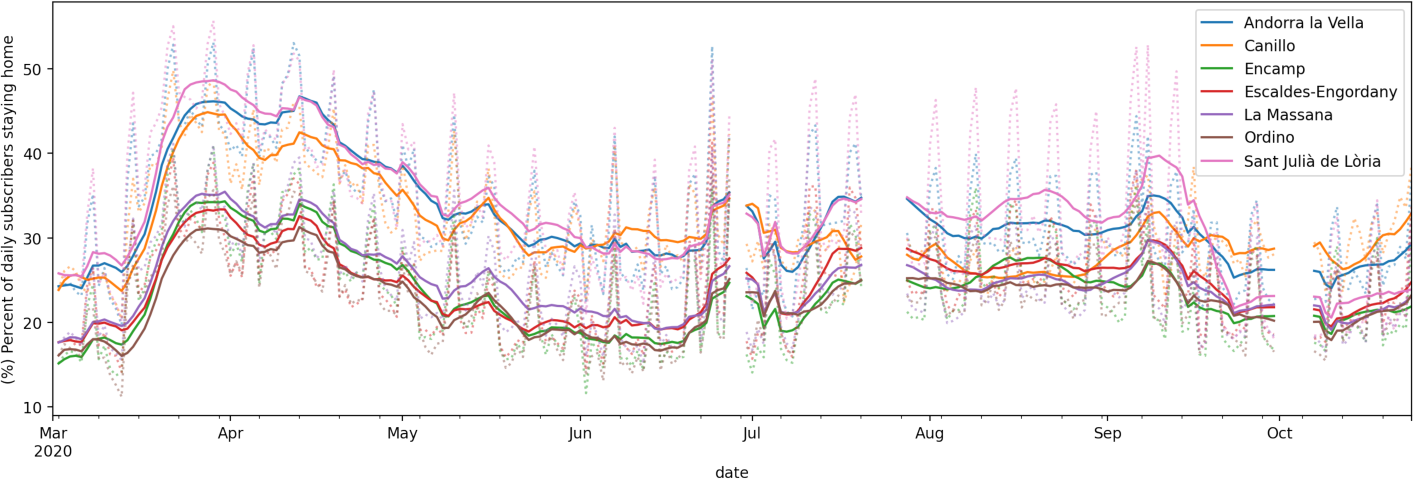
The daily percent of subscribers staying home by parish of residence. The dotted lines show daily estimates, while the solid lines show 7-day rolling averages.

The above described stay-at-home rate is normalised by parish population. In general, without normalisation, the mobility metrics are dominated by the most populous parishes, such as Andorra la Vella and Escaldes-Engordany. For example, the most populous parishes see the most trips in and out of them, as shown in Figure 9. The trends in mobility behaviour differed across parishes, possibly due to differences in tourism or other economic factors.

**Figure 9:**
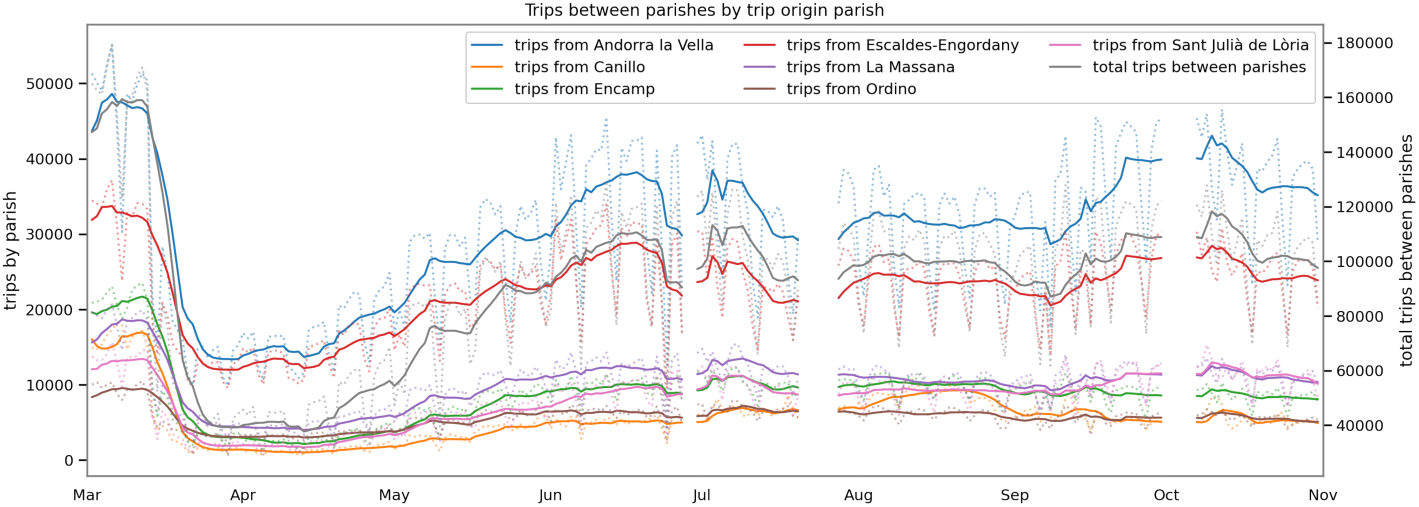
The daily number of trips between parishes, by parish origin. The dotted lines show daily estimates, while the solid lines show 7-day rolling averages.

#### 4.2.4 Distributions of Interaction Potential

The distributions of mobility behaviours were analyzed in order to determine the extent to which potentially risky mobility behaviours were dominated by relatively small numbers of people and events. This would be the case if the distributions of the mobility metrics had high positive skew or “fat-tails”. This concept is closely related to the Pareto Principle or 80/20 rule, the observation that in many real-world phenomena, 80% of the consequences come from 20% of the causes [34]. Quantities such as the number of people a person comes into contact with in a day, or the number of interactions which occur at an event can vary widely and may show such positive skew.

In order to evaluate how evenly or unevenly interaction potential was distributed across people, and how this changed over time, the daily contact graphs described in section 3.3.2 were used. The distributions of Daily Contacts were evaluated for a sample of 3 days in each month from March 2020 to October 2020 (the first available Monday-Wednesday period) in order to smooth out unusual behaviours on any one day. As shown in Figure 10, the distribution of number of connections per individual had a high positive skew in every period considered. Contacts were highest in March (before the lockdown) and lowest in April. For each month, the individuals were then ranked by their Daily Contacts and the top 20% of users were removed from the daily interaction graphs. It was found that by removing these top 20% most active people in each month, between 78% (May) and 85% (August) of the connections were eliminated.

**Figure 10:**
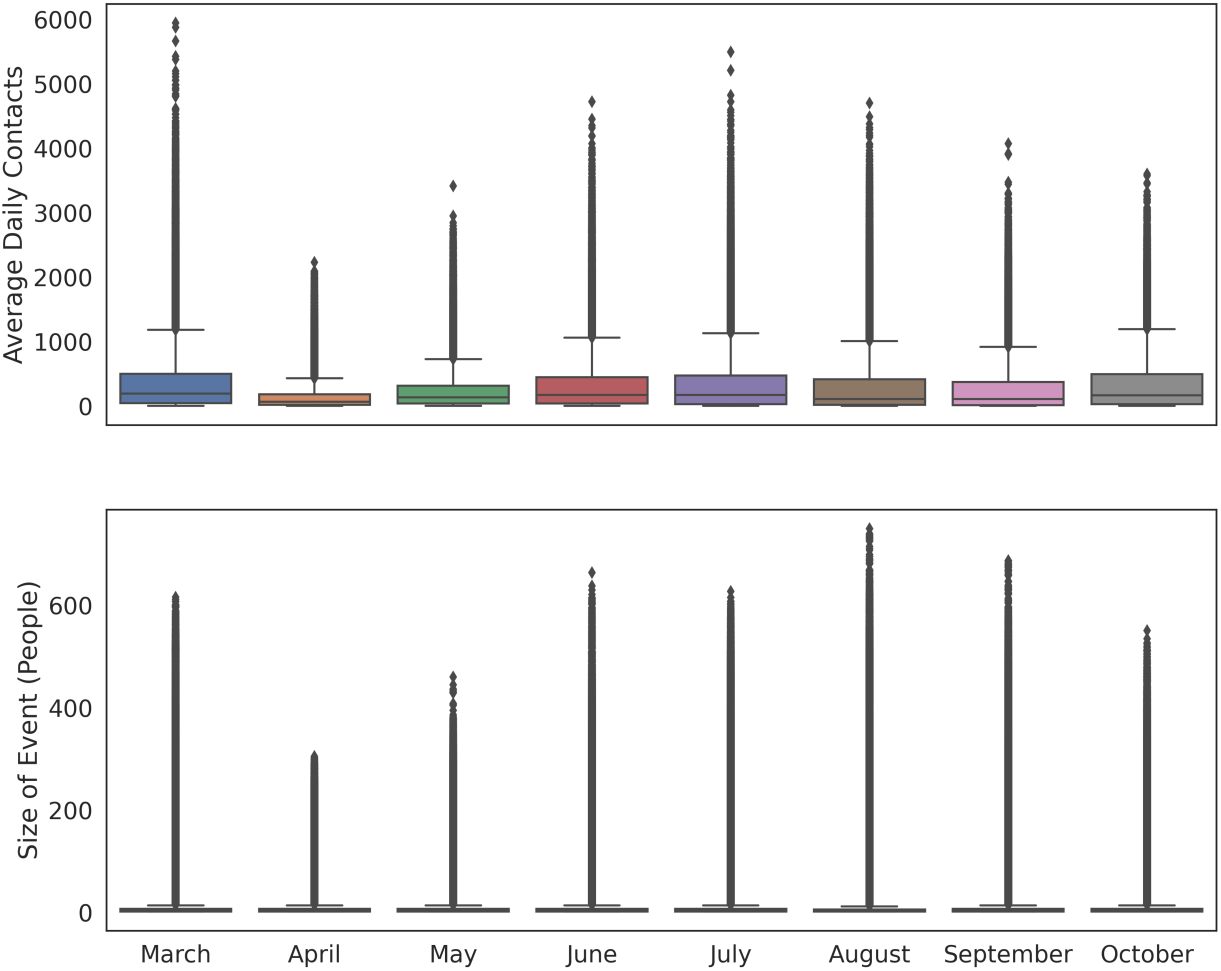
Box plots showing distributions of (Top) average daily contacts per person for 3-day periods and (Bottom) “event” sizes, in each month

In order to evaluate the contribution of potential super-spreader events, an event was defined as any spatio-temporal interval between March and October 2020 where at least 2 people were co-located, as defined in section 3.3.2. The distribution of events sizes was evaluated for each month. As shown in Figure 10, the number of people-per event showed high positive skew in each month. The distribution of event sizes contracted significantly in April and to a lesser extend in May and October. The interaction potential, *IP*_*tj*_, for every event was computed as described in section 3.3.2 and it was found that the top 20% of events across the whole period were responsible for 96% of all interaction potential.

### 4.3 Mobility and Transmission Rate

#### 4.3.1 Transmission Rate

The transmission rate of an infectious disease, *(3* can be simply defined as the average number of people an infected person would infect per unit time, if everyone they met were susceptible [8]. This metric can serve as a decent proxy for risky behaviours in a population. In standard SIR modelling, the rate of occurrence of new cases is equal to the product of the transmission rate, the number of currently infectious people and the fraction of the population who are still susceptible [25].

Therefore, a simple proxy for the transmission rate per day can be estimated by normalising the new infections per day by the number of currently infectious people and by the fraction of the population who are susceptible. A common approach to estimate this metric is to take the log case growth over a suitable period, such as 2 weeks [37]. In this study, the availability of an estimated reporting factor and the number of people present in the country on each day allowed for normalisation by a real-time estimated susceptible fraction. Note that the assumption of a constant reporting rate may have led to an overestimation of the susceptible population in the latter part of the study period. The rest of this subsection describes the calculation of the susceptible-normalised log case growth.

In order to smooth out stochastic variations in case reporting, 14 day rolling averages of new case reports, c(t), and cumulative case reports, cc(t), were taken. The population present on any day, n(t), was estimated as described in section 3.3.1. A reporting rate, *ρ*, of 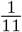 was estimated as described in section 4.1. The susceptible fraction of the population, S, was estimated as in equation 4:

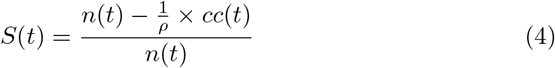

This estimate considers cumulative cases as the recovered population (‘R’ in SIR), with the naive assumption that infected people do not leave the country.

The susceptible-normalised new cases per day, C, were then estimated as in equation 5:

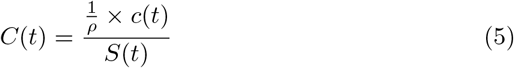

The following analysis uses the log of the growth in daily new cases, LCG, estimated as the relative change in the susceptible-normalised new cases since the 14 days prior (equation 6).

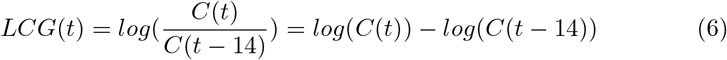

Figure 7.13 in the Supplementary Material shows a time series of the daily LCG(t) values along with the estimated daily values of the variables used in the calculation of LCG. Due to many days of zero cases between March and May, the LCG shows some volatility and undefined values.

#### 4.3.2 Mobility Metrics

Eight mobility metrics, shown in Figure 11, were compared to the LCG(t) over time. The mobility metrics all follow a broadly similar trend over time. They drop off sharply during March and remain close to their minimum levels throughout April. During May, the mobility metrics gradually increase, corresponding with a gradual relaxation of policies, or adherence to those policies. The five metrics related to trip making in Figure 11 ((i) the number of subscribers making trips between parishes, (ii) the number of trips between parishes, (iii) the portion of subscribers not staying home, (iv) the number of subscribers not staying home, (v) total trips) remain relatively steady after June at levels slightly lower than their peak in March. The indoor and outdoor interaction potential in Figure 11 appear to track one another very closely throughout the period of March to mid-September but after mid-September, these apparent trends diverge as indoor interactions increase while outdoor interactions decrease. This is likely due to crowded events moving indoors during the colder months. Entrances to the country, shown in Figure 11 increase throughout the summer, peaking in late August before decreasing.

**Figure 11:**
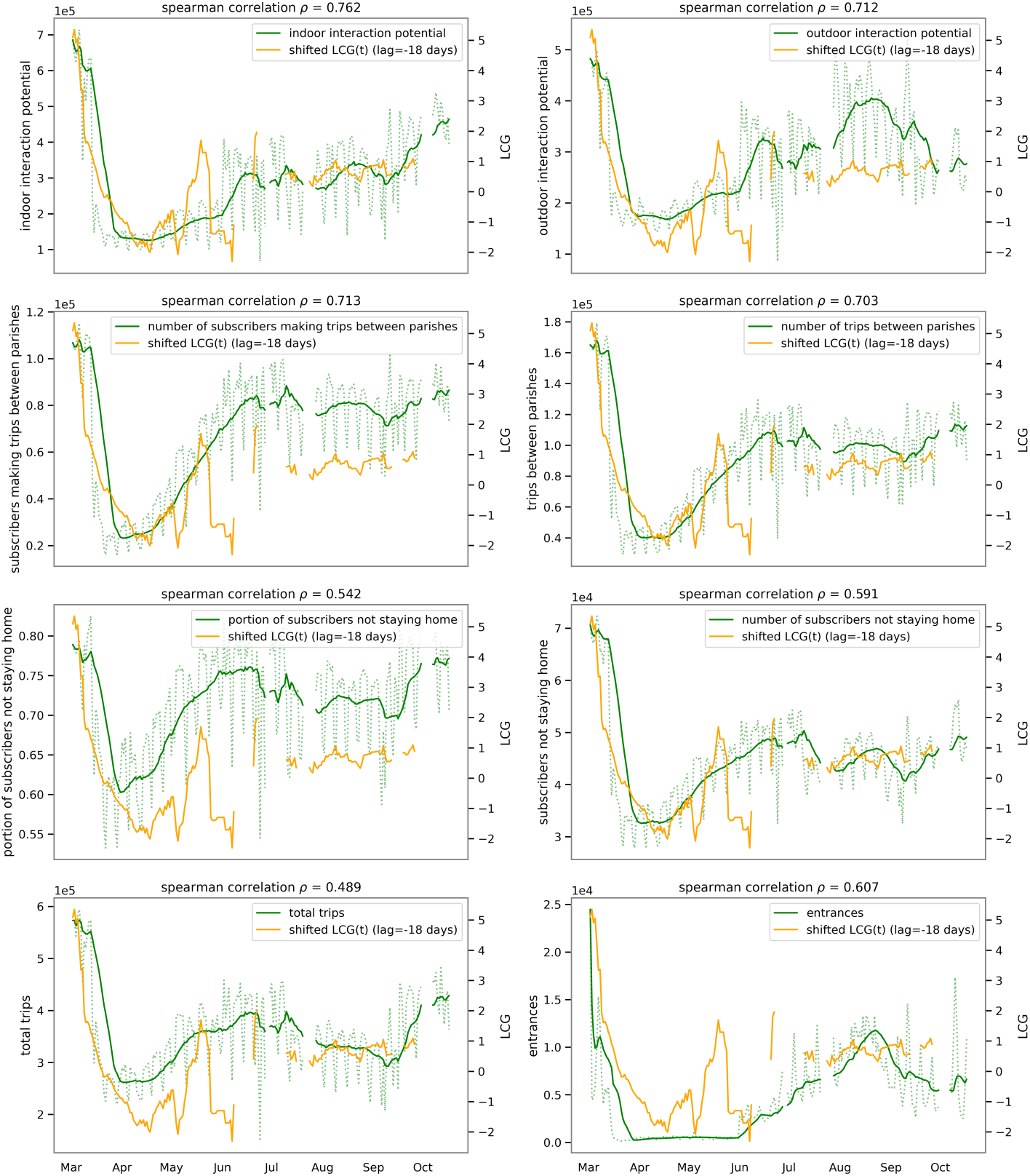
The time series for 8 mobility metrics are plotted ((i) indoor interaction potential, (ii) outdoor interaction potential, (iii) number of subscribers making trips between parishes, (iv) total trips between parishes, (v) portion of subscribers not staying home normalised by population, (vi) number of subscribers not staying home, (vii) total trips, (viii) country entrances). The time series for the estimated log case growth (LCG), shifted by a lag of -18 days is superimposed on the mobility metrics plots to better display the correlations between mobility and transmission rates. Changes in LCG are expected to lag behind changes in the mobility behaviors that affect transmission rate. The most notable correlation is between LCG and the indoor interaction potential metric (i).

#### 4.3.3 Correlations between transmission rate and mobility metrics

For any mobility metrics that are correlated with transmission rate, a lag should be expected between changes in these mobility metrics and changes in case growth. If mobility behaviours are responsible for infections, then the mobility metrics would be expected to lead infections by some period. The incubation period for COVID-19 can range up to 14 days [27] and there may be further delays between symptom onset, testing and reporting of results. On the other hand, if people restrict their mobility behaviours in response to changes in the rate of reported infections, then the infections would lead the mobility metrics by some period. For each metric, it is unknown a priori whether mobility leads infections, infections lead mobility, or both. In order to address this question, the time series data for daily mobility metrics were compared to the daily LCG time series with a range of lag times. The lag times ranged from -30 to 30 days, where a lag of -30 days corresponds with the mobility metrics leading the LCG time series by 30 days, and a lag of 30 days corresponds with the LCG leading the mobility metrics time series by 30 days. The Spearman’s Rank Correlation Coefficient was computed at every lag value. The Spearman’s Coefficient was deemed to be more appropriate than the Pearson’s Coefficient because it can identify monotonic relationships which are not necessarily linear.

As shown in Figure 12, several correlation patterns with transmission rate are present across the 8 mobility metrics considered. Figure 12 (A) shows 4 mobility metrics which are highly correlated with transmission rate, leading by 2-3 weeks. This suggests that when the behaviors associated with these mobility metrics change, transmission rates respond about 15-20 days later. Conversely, there is no evidence that these metrics increase or decrease in response to changes in transmission rate, as there is a small negative correlation between these mobility metrics and transmission rate when mobility metrics lag LCG. Figure 12 (B) shows 3 metrics which exhibit the same correlation trends as (A), but with slightly weaker correlation values. Again, these metrics are positively correlated with transmission with a lead of 2-3 weeks, and also moderately negatively correlated with transmission rate, with a lag of about 2-3 weeks. This suggests that (i) when these mobility behaviours increase, there is a moderate increase in transmission rate 2-3 weeks later and (ii) when the transmission rate increases, there is a moderate decrease in the mobility behaviours 2-3 weeks later. Figure 12 (C) shows that new entrances to the country seem to be highly correlated with infection rate, with a lead of 30 days. However, the correlation remains positive when changes in entrances lag LCG, making the relationship between country entrances and transmission less clear. Lead times greater than 30 days were not explored in this study because relationships between mobility and transmission rate at greater lead times would be less plausible.

**Figure 12:**
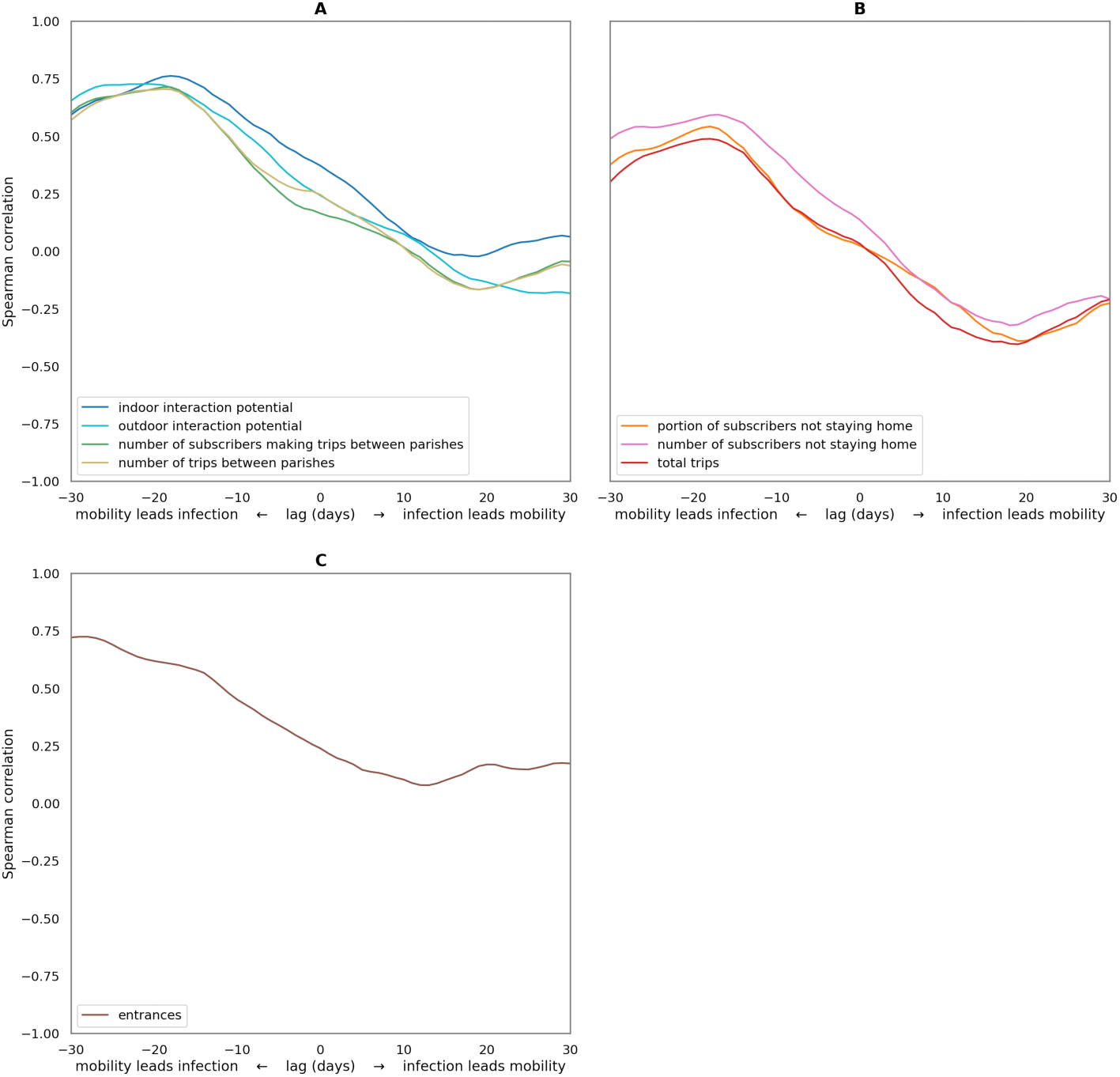
Correlations between the daily mobility metrics and log case growth (LCG) time series, for lags ranging from -30 to 30 days. Negative lag values correspond with the LCG time series shifted to lag behind the mobility metrics time series. e.g. At a lag of -30 days, mobility at day (t-30) is compared to LCG at day t, while a lag of (positive) 30 days measures the relationship between mobility metrics at day (t+30) and LCG at day t.

Table 3 in the Supplementary Material shows the numeric list of all correlation values between each mobility metric and LCG at each lag ranging from -30 to 30 days.

## 5 Discussion

The results of this analysis have highlighted a number of key insights about the nature of the spread of COVID-19 infection in Andorra. These insights and their potential policy implications are discussed in this section.

### 5.1 Tourism

Early in the COVID-19 pandemic, some of the most high-profile hotspots were European ski destinations and other areas which experienced high levels of international tourism. At the start of 2020, Andorra experienced a relatively normal ski season. This quickly transitioned in March to an economic lockdown, with border closures through the end of May. It therefore might have been expected that the areas with highest infection rates in May would be those where the most tourists had visited and interacted with residents before the lockdown. However, in this study it has been shown that there was no correlation between tourist contact and the COVID-19 exposure rates across Andorran parishes as measured in May 2020. In particular, the parish of Canillo accommodated 45% of tourists in February and residents of Canillo had a higher potential for interacting with tourists than residents of any other parish. Despite this, 3 of the 5 other parishes considered had an equal or higher infection rate. These findings suggest that, while tourists likely imported Andorra’s initial COVID-19 cases, subsequent geographical mixing quickly spread the infection to less touristic areas, even as the country was in full lockdown.

### 5.2 Social Distancing and Mobility Behaviour

Exposures to COVID-19 were distributed across the parishes of Andorra in a way that was unrelated to tourism or measured mobility behaviours. Possible reasons for this are that (i) the spatial resolution of data was coarse (7 parishes), there may have been unobserved socio-demographic differences between parishes which led to differences in infection rate or (iii) there was sufficient mixing across parish populations that the effects of differences in behavior did not last.

Changes in the infection transmission rate over time were closely related to changes in mobility behaviours of the population over time. Specifically, it was found that mobility metrics related to crowding and trips between parishes were highly correlated with infection rates 2-3 weeks later. Conversely, measures of overall trip making (including intra-parish travel) were only moderately correlated with later infection rates. Furthermore, the phenomenon most highly correlated with subsequent infection rates interaction potential - was shown to be a fat-tailed phenomenon dominated by a small number of people and events. Finally, some measures of stay-at-home rates and total trip making could be seen to reduce in response to high infection rates, which may have been due to policy responses or to individual volition.

These findings suggest that, in order to reduce transmission rates, policies should focus on encouraging people to remain distributed in their own home communities, thereby preventing both inter-community transmission and crowding. Furthermore, the events which draw the highest density of crowds should be identified and discouraged. On the other hand, policies which aim to reduce mobility across the board, are likely to be less effective and have higher social and economic costs. For example, between April 19th and May 13th in Andorra, outdoor exercise was restricted to a 1km radius for 1 hour every second day. Such measures would have likely reduced intra-parish outdoor trips while having little impact on the most relevant metrics like inter-parish trips and crowded indoor events.

The finding that trips between communities were correlated with transmission rate has important implications for simulating the transmission of respiratory diseases since many studies use areas significantly larger than the state of Andorra as the sub-populations. Also, the fact that interaction potential was highly correlated with transmission rate shows that the density with which people crowd together is an important determinant of transmission rate and therefore, the common assumption of mass-action (as discussed in section 1) is unrealistic for models of human respiratory disease spread.

### 5.3 Data Collection and Accuracy

The combination of population serology testing and telecoms data for all of Andorra provided a uniquely comprehensive dataset to study the progression of the pandemic in an entire country. However, the serology testing only occurred during a single period in May, 2020 and so the analysis of temporal trends in transmission rate were based on the public case report data. The fraction of true cases represented by these reports could not be precisely known. The combination of the serology data and case reports allowed estimation of this reporting factor for the period up to the start of May. However it is likely that the reporting rate subsequently changed with new testing procedures and policies, and as people entered and left the country. The problem of under-reporting is not unique to this study and in many studies there is no accounting for under-reporting as no serology data are available.

## 6 Conclusion

During the COVID-19 pandemic, policy makers worldwide were faced with the task of designing policies and interventions to try to curb the spread of infection. Numerous options have been available, including travel bans, economic shutdowns, stay-at-home orders and widespread testing to name a few. Yet all of these options are associated with social and economic costs and unknown levels of effectiveness. This study set out to provide decision support to policy-makers in Andorra and around the world by identifying which types of internal and external mobility are strongly associated with transmission risk and which are not, thereby enabling a greater focus on the most promising policies. A secondary contribution of this work was to provide empirical evidence to inform models for respiratory infection simulations.

The combination of a country-wide serology testing programme and high resolution telecoms data for the entire country presented a unique opportunity for granular insights into the relationship between mobility behaviour and COVID-19 exposure. Additionally, the isolation of Andorra during lockdown and the subsequent reintroduction of tourism provided a natural experiment to examine the relative importance of external versus internal mobility.

It has been shown in this study that internal mobility behaviours relating to crowding and rates of inter-community trips were most closely related to the spread of infection in Andorra. Conversely contact with tourists, intra-community trips and entrances to the country had lower or unclear correlations with transmission rates. Furthermore, interaction potential was characterised by highly skewed distributions with small numbers of people and events accounting for large proportions of the human contact.

Overall, the results of this study point policy-makers towards a targeted approach to mobility restrictions to curb the spread of COVID-19, based on discouraging the most risky behaviours rather than full economic lockdowns and border closures.

## Data Availability

The telecoms data and the serology screening data are not publicly available.

## Notes

* This work was supported by the Andorra Innovation Hub.

### Competing Interest Statement

The authors have declared no competing interest.

### Funding Statement

The Andorra Innovation Hub provided financial support to the City Science Group at MIT Media Lab

### Author Declarations

The serological screening was approved by the relevant Andorran regulatory agencies and the local Research Ethics Committee. Participation in the screening was entirely voluntary. The Institutional Review Board of the Servei Andorra Atencio Sanitaria (SAAS) approved the study (register number 0720). The study was supported and approved by the Andorran government.

### Summary of Updates

Spelling of author name revised. Added reference to the original serological screening study. Added details of Institutional Review Board authorisation for the original serological screening.

