## Supplemental Materials for "Mobility and COVID-19 in Andorra: Country-scale analysis of high-resolution mobility patterns and infection spread"

### 7 Supplementary Material

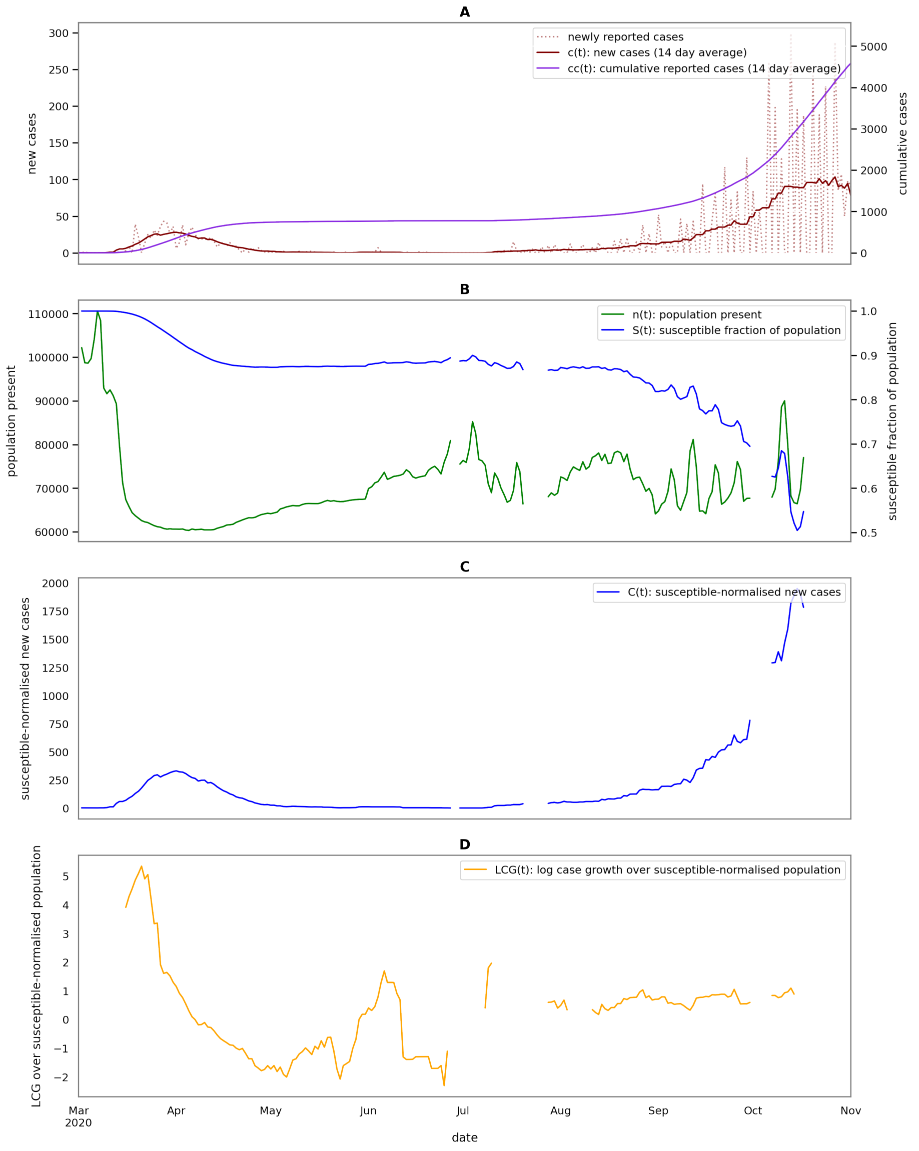


Figure 7.13: LCG(t) as well as variables used in the calculation, as described in section 4.3.1: (A) Newly reported cases (not smoothed), c(t): newly reported cases (14 day rolling average), cc(t): cumulative reported cases (14 day rolling average), (B) n(t): population present, S(t): susceptible fraction of the population, (C) C(t): The susceptible-normalised new cases per day, (D) LCG(t): log of the growth in daily susceptible-normalised new cases since the 14 days prior.

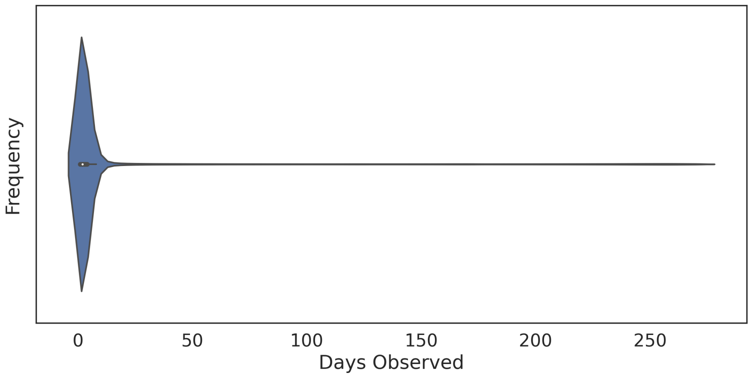


Figure 7.14: Distribution of number of days observed by subscriber. The distribution is dominated by tourists who significantly outnumber residents.

| lag | indoor interaction potential | outdoor interaction potential | people making trips between parishes | total trips between parishes | fraction of people not staying home | people making trips | total trips | country entrances |
|---|---|---|---|---|---|---|---|---|
| -30 | 0.592 | 0.653 | 0.602 | 0.57 | 0.376 | 0.489 | 0.301 | 0.721 |
| -29 | 0.619 | 0.679 | 0.631 | 0.598 | 0.405 | 0.513 | 0.338 | 0.724 |
| -28 | 0.637 | 0.698 | 0.65 | 0.624 | 0.425 | 0.529 | 0.367 | 0.724 |
| -27 | 0.655 | 0.714 | 0.663 | 0.644 | 0.438 | 0.541 | 0.394 | 0.718 |
| -26 | 0.664 | 0.722 | 0.67 | 0.659 | 0.441 | 0.541 | 0.414 | 0.707 |
| -25 | 0.671 | 0.723 | 0.674 | 0.67 | 0.446 | 0.538 | 0.425 | 0.69 |
| -24 | 0.682 | 0.723 | 0.68 | 0.68 | 0.458 | 0.54 | 0.436 | 0.67 |
| -23 | 0.696 | 0.726 | 0.688 | 0.691 | 0.475 | 0.547 | 0.449 | 0.653 |
| -22 | 0.712 | 0.726 | 0.693 | 0.697 | 0.491 | 0.554 | 0.461 | 0.637 |
| -21 | 0.731 | 0.727 | 0.699 | 0.7 | 0.508 | 0.563 | 0.47 | 0.627 |
| -20 | 0.746 | 0.726 | 0.706 | 0.701 | 0.524 | 0.573 | 0.48 | 0.619 |
| -19 | 0.758 | 0.723 | 0.713 | 0.705 | 0.537 | 0.583 | 0.487 | 0.613 |
| -18 | 0.762 | 0.712 | 0.713 | 0.703 | 0.542 | 0.591 | 0.489 | 0.607 |
| -17 | 0.759 | 0.699 | 0.701 | 0.693 | 0.533 | 0.594 | 0.483 | 0.601 |
| -16 | 0.747 | 0.681 | 0.672 | 0.668 | 0.508 | 0.585 | 0.469 | 0.59 |
| -15 | 0.729 | 0.658 | 0.639 | 0.637 | 0.476 | 0.571 | 0.448 | 0.58 |
| -14 | 0.711 | 0.636 | 0.613 | 0.613 | 0.449 | 0.557 | 0.429 | 0.567 |
| -13 | 0.682 | 0.608 | 0.572 | 0.574 | 0.402 | 0.527 | 0.387 | 0.54 |
| -12 | 0.66 | 0.588 | 0.532 | 0.534 | 0.363 | 0.493 | 0.344 | 0.509 |
| -11 | 0.638 | 0.57 | 0.493 | 0.499 | 0.326 | 0.458 | 0.309 | 0.478 |
| -10 | 0.605 | 0.54 | 0.448 | 0.454 | 0.271 | 0.428 | 0.266 | 0.451 |
| -9 | 0.574 | 0.509 | 0.404 | 0.413 | 0.223 | 0.397 | 0.225 | 0.429 |
| -8 | 0.548 | 0.482 | 0.362 | 0.378 | 0.185 | 0.358 | 0.187 | 0.407 |
| -7 | 0.53 | 0.45 | 0.328 | 0.351 | 0.161 | 0.323 | 0.168 | 0.381 |
| -6 | 0.509 | 0.415 | 0.293 | 0.327 | 0.132 | 0.29 | 0.141 | 0.359 |
| -5 | 0.475 | 0.374 | 0.259 | 0.304 | 0.101 | 0.258 | 0.116 | 0.34 |
| -4 | 0.451 | 0.34 | 0.228 | 0.285 | 0.079 | 0.231 | 0.098 | 0.319 |
| -3 | 0.432 | 0.311 | 0.203 | 0.271 | 0.065 | 0.205 | 0.083 | 0.296 |
| -2 | 0.408 | 0.284 | 0.186 | 0.263 | 0.049 | 0.182 | 0.065 | 0.277 |
| -1 | 0.391 | 0.263 | 0.179 | 0.26 | 0.04 | 0.162 | 0.053 | 0.256 |
| 0 | 0.371 | 0.242 | 0.164 | 0.245 | 0.024 | 0.136 | 0.033 | 0.239 |
| 1 | 0.346 | 0.22 | 0.152 | 0.22 | 0.006 | 0.102 | 0.002 | 0.216 |
| 2 | 0.324 | 0.199 | 0.144 | 0.197 | -0.012 | 0.069 | -0.023 | 0.196 |
| 3 | 0.301 | 0.178 | 0.134 | 0.18 | -0.031 | 0.036 | -0.053 | 0.184 |
| 4 | 0.274 | 0.157 | 0.12 | 0.161 | -0.051 | -0.009 | -0.097 | 0.168 |
| 5 | 0.242 | 0.144 | 0.104 | 0.138 | -0.074 | -0.052 | -0.142 | 0.146 |
| 6 | 0.21 | 0.128 | 0.09 | 0.116 | -0.098 | -0.088 | -0.185 | 0.137 |
| 7 | 0.177 | 0.113 | 0.076 | 0.094 | -0.117 | -0.116 | -0.219 | 0.132 |
| 8 | 0.142 | 0.099 | 0.058 | 0.068 | -0.137 | -0.143 | -0.247 | 0.124 |
| 9 | 0.118 | 0.088 | 0.041 | 0.047 | -0.158 | -0.165 | -0.268 | 0.112 |
| 10 | 0.087 | 0.074 | 0.018 | 0.015 | -0.192 | -0.196 | -0.302 | 0.103 |
| 11 | 0.059 | 0.054 | -0.007 | -0.018 | -0.224 | -0.223 | -0.331 | 0.088 |
| 12 | 0.044 | 0.033 | -0.027 | -0.04 | -0.24 | -0.236 | -0.34 | 0.079 |
| 13 | 0.022 | 0.004 | -0.059 | -0.073 | -0.272 | -0.259 | -0.358 | 0.079 |
| 14 | 0.006 | -0.022 | -0.085 | -0.099 | -0.305 | -0.28 | -0.374 | 0.087 |
| 15 | -0.007 | -0.051 | -0.11 | -0.119 | -0.332 | -0.295 | -0.384 | 0.1 |
| 16 | -0.016 | -0.081 | -0.132 | -0.139 | -0.355 | -0.304 | -0.393 | 0.113 |
| 17 | -0.015 | -0.102 | -0.147 | -0.151 | -0.361 | -0.309 | -0.392 | 0.125 |
| 18 | -0.021 | -0.12 | -0.162 | -0.164 | -0.376 | -0.322 | -0.402 | 0.144 |
| 19 | -0.022 | -0.126 | -0.167 | -0.167 | -0.39 | -0.319 | -0.405 | 0.162 |
| 20 | -0.014 | -0.134 | -0.161 | -0.163 | -0.389 | -0.303 | -0.394 | 0.169 |
| 21 | 0.0 | -0.145 | -0.154 | -0.155 | -0.378 | -0.284 | -0.374 | 0.168 |
| 22 | 0.016 | -0.153 | -0.141 | -0.141 | -0.362 | -0.269 | -0.355 | 0.158 |
| 23 | 0.03 | -0.163 | -0.13 | -0.128 | -0.351 | -0.258 | -0.338 | 0.151 |
| 24 | 0.039 | -0.173 | -0.114 | -0.118 | -0.339 | -0.243 | -0.321 | 0.148 |
| 25 | 0.042 | -0.18 | -0.102 | -0.107 | -0.325 | -0.226 | -0.301 | 0.147 |
| 26 | 0.047 | -0.181 | -0.09 | -0.098 | -0.313 | -0.219 | -0.287 | 0.155 |
| 27 | 0.056 | -0.182 | -0.073 | -0.081 | -0.284 | -0.208 | -0.262 | 0.163 |
| 28 | 0.063 | -0.178 | -0.057 | -0.067 | -0.256 | -0.201 | -0.24 | 0.173 |
| 29 | 0.068 | -0.178 | -0.043 | -0.057 | -0.234 | -0.194 | -0.219 | 0.175 |
| 30 | 0.063 | -0.183 | -0.045 | -0.063 | -0.226 | -0.208 | -0.21 | 0.173 |

Table 3: The correlations between the daily time series of 8 mobility metrics and lagged log case growth (LCG). LCG is shifted by lags ranging from -30 to 30 days.
